# Obesity-Related Metabolites are Associated with Incident Coronary Heart Disease and Respond to Metabolic and Bariatric Surgery

**DOI:** 10.64898/2026.03.06.26347826

**Authors:** Zicheng Wang, Yulu Zheng, Lei Wang, Charles R. Flynn, Xiao-Ou Shu, Qiuyin Cai, Deepak K. Gupta, Loren Lipworth, Wei Zheng, Xinmeng Zhang, You Chen, Jason M. Samuels, Danxia Yu

**Author notes:** **Correspondence:** Danxia Yu, PhD, 2525 West End Avenue, Nashville, TN, USA 37203.

## Abstract

**Objective:** Obesity is a major risk factor for coronary heart disease (CHD). This study aims to develop a metabolite signature of body mass index (BMI-MetSig) then assess its association with incident CHD and responsiveness to metabolic and bariatric surgery (MBS).

**Research Design and Methods:** In a case-control study of incident CHD nested within the Southern Community Cohort Study (SCCS) including 600 case-control pairs, we used elastic net regression with 10-fold cross-validation to derive the BMI-MetSig. Associations of BMI-MetSig with incident CHD was examined using conditional logistic regression in the nested case-control study. Further, in a cohort of 95 patients who received MBS, we evaluated this BMI-MetSig in association with estimated 30-year cardiovascular disease (CVD) risks, which was estimated by the American Heart Association’s PREVENT equations, and examined changes of its constituent metabolites after surgery using linear mixed-effects models.

**Results:** In the SCCS, the BMI-MetSig, comprising 94 metabolites, was significantly associated with incident CHD risk among all participants (OR per standard deviation [SD] increase: 1.48; 95% CI, 1.28–1.71) and across subgroups. Among MBS patients, the BMI-MetSig was significantly associated with increased estimated 30-year risks of CHD (β per SD increase: 1.29; p<0.001) and other CVDs. Levels of 17 (20.0%) and 19 (22.4%) metabolites in the BMI-MetSig significantly changed 3- and 12-month post-surgery (FDR<0.10 and log_2_FC > 0.15), including choline and acetyl-2-aminoadipate.

**Conclusions:** The BMI-MetSig is associated with higher CHD incidence and estimated 30-year CVD risks and responds to MBS. BMI-MetSig may serve as a blood-based biomarker for cardiometabolic risk stratification and monitoring.

## INTRODUCTION

Coronary heart disease (CHD), a specific type of atherosclerosis cardiovascular disease (ASCVD) where plaque builds up in the coronary arteries, remains a leading cause of death in the United States despite decades of progress in its prevention and treatment. In 2022, approximately one in twenty American adults aged 20 years or older had CHD, and over 371,000 Americans died from this disease^1^. The lifetime risk of CHD is 48.6% for men and 31.7% for women at age 40 years^2^. Chronic inflammation, endothelial dysfunction, and plaque formation and progression are key biological mechanisms underlying CHD pathogenesis^3^. Obesity is a major modifiable risk factor that contributes to these pathways and significantly elevates the risk of CHD and related cardiovascular outcomes^4–6^. Metabolic and bariatric surgery (MBS) is one of the most effective and durable interventions to treat morbid obesity.

In recent years, metabolomics—the high-throughput profiling of small-molecule metabolites—has emerged as a promising tool for uncovering the biochemical underpinnings of cardiometabolic diseases^7^. Several studies have applied metabolomics to identify metabolite predictors of CHD risk, highlighting lipids (e.g., phosphatidylcholines, lysoPCs, sphingolipids), amino acids, and other bioactive molecules^8–11^. Parallel work in obesity research has identified metabolite alterations reflecting insulin resistance, lipid oxidation, gut microbial metabolism (e.g., trimethylamine N-oxide, TMAO), and hormonal dysregulation^12–15^. For example, a 2019 systematic review summarized findings from 60 obesity-focused metabolomics studies and reported consistent associations of obesity with branched-chain and aromatic amino acids, acylcarnitines, and certain lipid metabolites^16^.

While obesity and CHD have each been extensively studied independently using metabolomics, few studies have investigated metabolites and metabolic pathways linking obesity to CHD risk. Meanwhile, studies have evaluated how obesity-related metabolites change after clinical weight-loss interventions is scarce. Without assessing modifiability, the biomarker and therapeutic potentials of identified metabolites remain limited. Therefore, the knowledge gap lies in understanding the specific metabolite signatures that underpin the relationship between obesity and CHD across diverse populations, and how these obesity-related metabolites could be altered by clinical interventions, including with highly effective MBS.

In this study, we aim to fill this gap by integrating data from two sources. The first is a large, diverse, nested case-control study within the Southern Community Cohort Study (SCCS), with half of the participants self-identifying as Black or African American. The second is a longitudinal study of patients who underwent MBS (the Gut Microbiota in Metabolic Surgery Study [GUMMY]). We aim to identify metabolites and molecular pathways linking obesity to CHD risk and inform future precision prevention strategies by: (1) developing a metabolite signature of body mass index (BMI-MetSig) using an untargeted profiling panel of metabolites in SCCS baseline blood samples; (2) evaluating the association between BMI-MetSig and incident CHD in SCCS; (3) evaluating how this BMI-MetSig and its constituent metabolites may be modified by MBS and whether the BMI-MetSig is related to post-surgery cardiometabolic improvements in GUMMY.

## METHODS

### Study Cohorts

The SCCS is a prospective cohort of ∼85,000 predominantly low-income individuals living in the southeastern U.S., enrolled from community health centers between 2002 and 2009^17^. A nested case-control study included 600 incident CHD cases and 600 age-/sex-/race-matched controls (300 pairs per race: Black/African American and White) selected based on following inclusion criteria: absence of baseline cardiovascular diseases (CVD) and availability of baseline plasma and the Centers for Medicare and Medicaid Services (CMS) claims data for CHD ascertainment^18^. After excluding participants with missing metabolomics data (n=6), missing baseline BMI or had extreme values (BMI ≤ 18.5 or BMI ≥ 52.06) (n=24), 1,170 participants were included in the analysis.

The GUMMY is an ongoing longitudinal study of patients aged 21–65 undergoing primary Roux-en-Y gastric bypass (RYGB) or sleeve gastrectomy (SG) at Vanderbilt University Medical Center (VUMC). Inclusion required one or more metabolic conditions (prediabetes or diabetes, hypertension, and dyslipidemia) and written consent to provide questionnaire answers and biospecimens. Patients were excluded if they had gastrointestinal disease (inflammatory bowel disease and celiac disease), history of major CVDs (CHD, stroke, and heart failure [HF]), chemo/radiation therapy for cancer within 5 years, antibiotic use within 3 months, or gastrointestinal disturbances (vomiting, constipation, and diarrhea) within 7 days before enrollment. This ongoing study has so far included 95 patients with clinical and metabolomics data at pre-surgery baseline (T0), and 3 months (T3) and 12 months (T12) post-surgery. A graphical representation of our study design is shown in **Figure S1**.

The SCCS and the GUMMY were both approved by the Institutional Review Board of VUMC. Reporting of the results adhered to the Strengthening of the Reporting of Observational Studies in Epidemiology guidelines^19^.

### Outcomes, Exposure, and Covariates

In SCCS, CHD cases were identified through CMS claims using validated codes and through the National Death Index, with CHD listed as the underlying cause of death. Plasma lipoprotein profiles, including triglycerides (TG), high-density lipoprotein cholesterol (HDL-C), and non-HDL-C were measured via nuclear magnetic resonance (NMR) using the Bruker IVDr platform at the Vanderbilt NMR Core Facility. BMI and covariates were collected via structured questionnaires at baseline. Sociodemographic variables include age, sex, self-reported race (White, Black/African American), education level (less than high school, completed high school, beyond completion of high school), and annual family income (<$15000, ≥$15000). Lifestyle factors are smoking status (current, former, never), alcohol drinks per day as a continuous variable, total physical activity measured in metabolic equivalents hours per week, and Healthy Eating Index per *Dietary Guidelines for Americans* 2010^20^. Medical history considered self-reported diagnosis or medication use for diabetes, hypertension, and dyslipidemia.

In GUMMY, plasma lipoproteins and creatinine were quantified using the Nightingale Health NMR biomarker platform (Helsinki, Finland), and estimated glomerular filtration rate (eGFR) was then calculated based on creatinine concentration^21^. We estimated the thirty-year risk scores (0-100%) for CHD, ASCVD, total CVD, and HF following the American Heart Association’s PREVENT equations^22^. Variables used to evaluate 30-year risks for all outcomes include age, systolic blood pressure, diabetes status, current smoking status, eGFR, and anti-hypertensive use. The models for total CVD, ASCVD, and CHD additionally included HDL-C, non-HDL-C, and statin use, which the model for HF additionally included BMI. These variables were either extracted from electronic health records (EHRs) or collected from questionnaires at each clinical visit. BMI at T0, T3, and T12 were measured at the VUMC Weight Loss Center. Sociodemographic (age, sex, and self-reported race [Black/African American, White, other]) and clinical data (operation type [SG, RYGB] and histories of diabetes, hypertension, and dyslipidemia by diagnosis or medication use) were extracted from EHRs.

### Metabolomics Profiling

All plasma samples were profiled by untargeted LC-MS/MS (Metabolon Inc.) following a standard protocol^23^. Briefly, baseline plasma from matched CHD case–control pairs were retrieved and placed adjacently in the same assay batch while laboratory staff were blinded to case–control status. For the GUMMY, each patient’s pre- and post-operative samples were likewise handled. This handling helped minimize potential batch effects. Plasma was extracted with methanol and divided into four aliquots for analyses in positive and negative ion modes using both reverse-phase and HILIC chromatography. Metabolites were called by automated matching of spectral features to a reference library of >4,000 authenticated standards, followed by visual review for quality control. Most metabolites (>80%) were annotated against internal standards, and those identified only by a match to a known MS spectrum or to a chemical formula were flagged with ‘*’ and ‘**’, respectively. Peak areas were used for quantification. In total, 1,502 metabolites were detected in the SCCS and 1,602 in the GUMMY. After excluding metabolites with >80% missingness across participants in the SCCS and >50% missingness across all three time points in the GUMMY, 1,478 and 1,199 metabolites remained, respectively. Missing values were imputed as one-half of the minimum observed value among non-missing samples. All metabolite values were log-transformed and standardized. Additional details can be found in our prior publications^9,18,24^.

### Statistical Analysis

In the SCCS, we first tested BMI–metabolite associations using multivariable linear models (overall and sex-stratified), keeping metabolites with a false discovery rate (FDR) < 0.10. With those metabolites, we then fit an elastic-net model with 10-fold cross-validation to derive the BMI-MetSig. We tested the proportion of BMI variance explained by the signature through fitting a simple linear regression of BMI on the BMI-MetSig. After removing participants not in CHD case-control pairs (n=24), we used covariate-adjusted conditional logistic regression models to assess associations of each standard deviation (1-SD) increase in BMI and BMI-MetSig with incident CHD in the remaining 1,146 participants (573 pairs). Linear regression models were used to examine the associations of BMI and BMI-MetSig with baseline lipoprotein levels. Stratified analysis was performed by sex and race using conditional logistic regression as well as smoking status, BMI category, and histories of hypertension, diabetes, and dyslipidemia using unconditional logistic regression. Interaction terms were evaluated by the Wald test. To assess the robustness of the BMI-MetSig, we conducted a leave-one-out sensitivity analysis, iteratively removing each metabolite and reassessing its association with CHD. In addition, each metabolite within the BMI-MetSig was individually tested for association with incident CHD with FDR adjustment. All models adjusted for education level, annual family income, and lifestyle covariates when in strata of matching pairs, and additionally age, sex, and race when in strata where matching pairs were broken.

In GUMMY, BMI-MetSig were calculated at T0, T3, and T12 using weights of the 85 available metabolites derived from the SCCS. Associations of BMI-MetSig with non-HDL-C, HDL-C, TG, eGFR, and estimated 30-year CVD risk scores were assessed using linear mixed-effects models. Individual metabolite changes over time were evaluated using mixed-effects models and fold-change analysis. Metabolites were considered significantly altered if they exhibited absolute log₂ fold changes >0.15 (approximately 11% change) and FDR <0.10, with the direction of change consistent with the BMI-MetSig weight (i.e., metabolites with positive weights decreased and those with negative weights increased after MBS). Models adjusted for age, sex, and self-reported operation type, and histories of diabetes, hypertension, and dyslipidemia. All analyses were performed in R (*v4.2.2*).

## RESULTS

### Participant Characteristics

**Table 1** presents baseline characteristics of participants in the SCCS CHD case-control study by baseline obesity status and incident CHD status. The mean age was 55 years (SD ∼8.5). The mean BMI was approximately 25.0 kg/m² (SD ∼2.9) in the non-obese group and 36.0 kg/m² (SD ∼4.9) in the obese group. Regardless of obesity status, incident CHD cases had lower levels of educational attainment and household income, were more likely to be current or former smokers, and had lower diet quality and higher prevalence of diabetes and dyslipidemia. The prevalence of diabetes, hypertension, and dyslipidemia was 44.3%, 73.0%, and 47.6% among incident CHD cases with obesity at baseline. **Table S1** presents the baseline characteristics of 95 GUMMY participants. The mean BMI was 45.5 kg/m² (SD: 6.11), and the mean age was 44.9 years (SD: 9.57). The majority were female (81.1%), had a history of diabetes (60.0%), hypertension (83.2%), and dyslipidemia (56.8%).

**Table 1.**
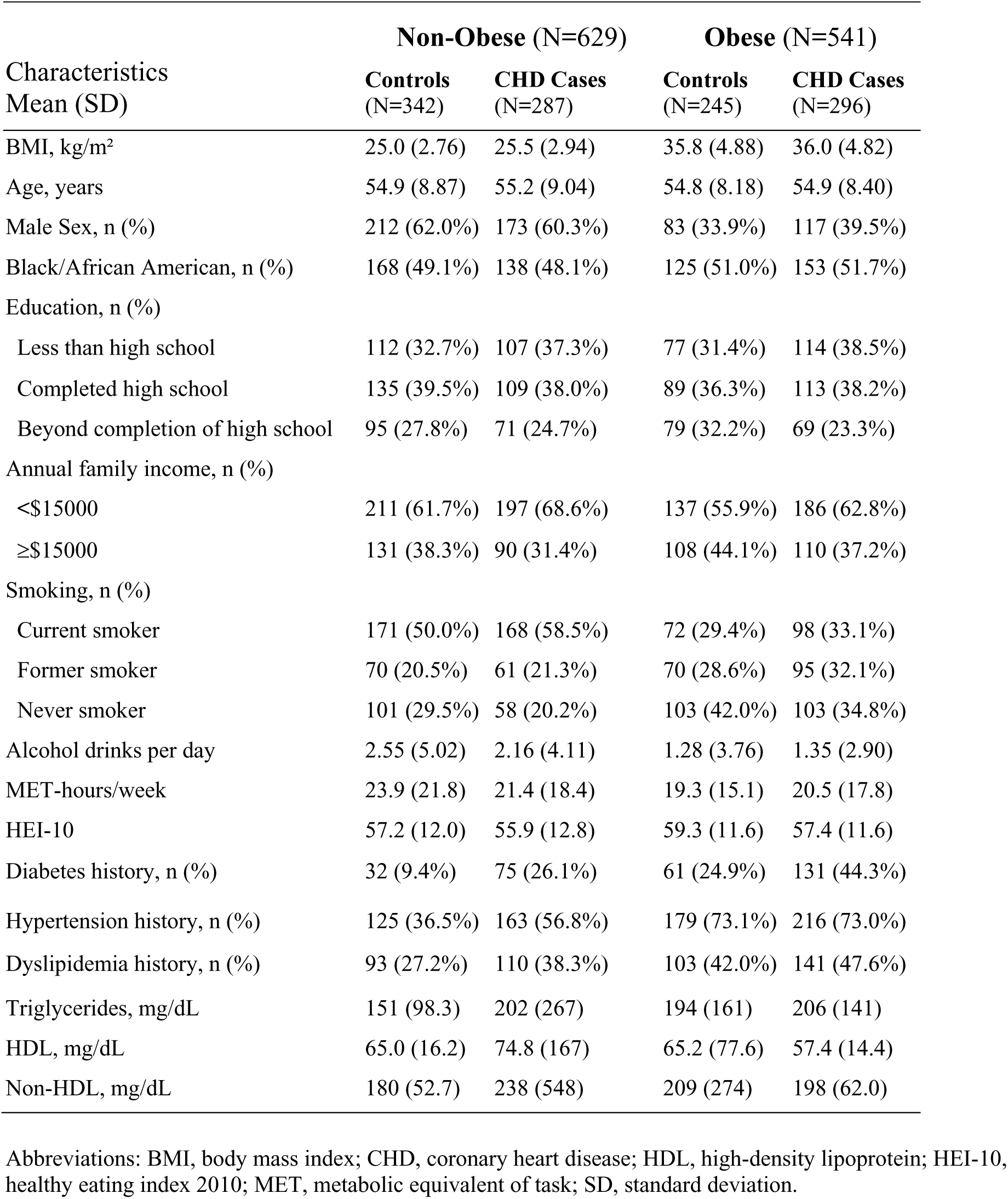
Baseline characteristics of participants in the nested case-control study within the Southern Community Cohort Study (SCCS)

| Characteristics<br>Mean (SD) | Non-Obese (N=629) |  | Obese (N=541) |  |
| --- | --- | --- | --- | --- |
|  | Controls<br>(N=342) | CHD Cases<br>(N=287) | Controls<br>(N=245) | CHD Cases<br>(N=296) |
| BMI, kg/m<sup>2</sup> | 25.0 (2.76) | 25.5 (2.94) | 35.8 (4.88) | 36.0 (4.82) |
| Age, years | 54.9 (8.87) | 55.2 (9.04) | 54.8 (8.18) | 54.9 (8.40) |
| Male Sex, n (%) | 212 (62.0%) | 173 (60.3%) | 83 (33.9%) | 117 (39.5%) |
| Black/African American, n (%) | 168 (49.1%) | 138 (48.1%) | 125 (51.0%) | 153 (51.7%) |
| Education, n (%) |  |  |  |  |
| Less than high school | 112 (32.7%) | 107 (37.3%) | 77 (31.4%) | 114 (38.5%) |
| Completed high school | 135 (39.5%) | 109 (38.0%) | 89 (36.3%) | 113 (38.2%) |
| Beyond completion of high school | 95 (27.8%) | 71 (24.7%) | 79 (32.2%) | 69 (23.3%) |
| Annual family income, n (%) |  |  |  |  |
| <\$15000 | 211 (61.7%) | 197 (68.6%) | 137 (55.9%) | 186 (62.8%) |
| ≥\$15000 | 131 (38.3%) | 90 (31.4%) | 108 (44.1%) | 110 (37.2%) |
| Smoking, n (%) |  |  |  |  |
| Current smoker | 171 (50.0%) | 168 (58.5%) | 72 (29.4%) | 98 (33.1%) |
| Former smoker | 70 (20.5%) | 61 (21.3%) | 70 (28.6%) | 95 (32.1%) |
| Never smoker | 101 (29.5%) | 58 (20.2%) | 103 (42.0%) | 103 (34.8%) |
| Alcohol drinks per day | 2.55 (5.02) | 2.16 (4.11) | 1.28 (3.76) | 1.35 (2.90) |
| MET-hours/week | 23.9 (21.8) | 21.4 (18.4) | 19.3 (15.1) | 20.5 (17.8) |
| HEI-10 | 57.2 (12.0) | 55.9 (12.8) | 59.3 (11.6) | 57.4 (11.6) |
| Diabetes history, n (%) | 32 (9.4%) | 75 (26.1%) | 61 (24.9%) | 131 (44.3%) |
| Hypertension history, n (%) | 125 (36.5%) | 163 (56.8%) | 179 (73.1%) | 216 (73.0%) |
| Dyslipidemia history, n (%) | 93 (27.2%) | 110 (38.3%) | 103 (42.0%) | 141 (47.6%) |
| Triglycerides, mg/dL | 151 (98.3) | 202 (267) | 194 (161) | 206 (141) |
| HDL, mg/dL | 65.0 (16.2) | 74.8 (167) | 65.2 (77.6) | 57.4 (14.4) |
| Non-HDL, mg/dL | 180 (52.7) | 238 (548) | 209 (274) | 198 (62.0) |

Abbreviations: BMI, body mass index; CHD, coronary heart disease; HDL, high-density lipoprotein; HEI-10, healthy eating index 2010; MET, metabolic equivalent of task; SD, standard deviation.

### BMI-MetSig Identification

Among the participants, 265 metabolites were significantly associated with BMI (FDR < 0.1). Elastic-net regression selected 94 metabolites to compose the BMI-MetSig (**Figure 1a**), which explained 60.32% of the BMI variance. Most of these metabolites were from the super pathways of lipids (n = 40) and amino acids (n = 16). The top five metabolites with negative weights were four lipids (3beta-hydroxy-5-cholestenoate, 1-lignoceroyl-GPC [24:0], palmitoyl sphingomyelin [d18:1/16:0], and 1-myristoyl-2-arachidonoyl-GPC [14:0/20:4]) and an amino acid metabolite (3-hydroxy-2-ethylpropionate). The top five positively associated metabolites include four lipid metabolites (two sphingomyelins, glycerol, and cortolone glucuronide [1]) and one unknown compound. The BMI-MetSig was highly correlated with BMI among all participants (Spearman’s correlation coefficient [R] = 0.79), sex-specific groups (R = 0.75 in women; R = 0.79 in men) (**Figure 1b**), and race-specific groups (R = 0.79 in Black; R = 0.80 in White participants) (**Figure 1c**).

**Fig. 1.**
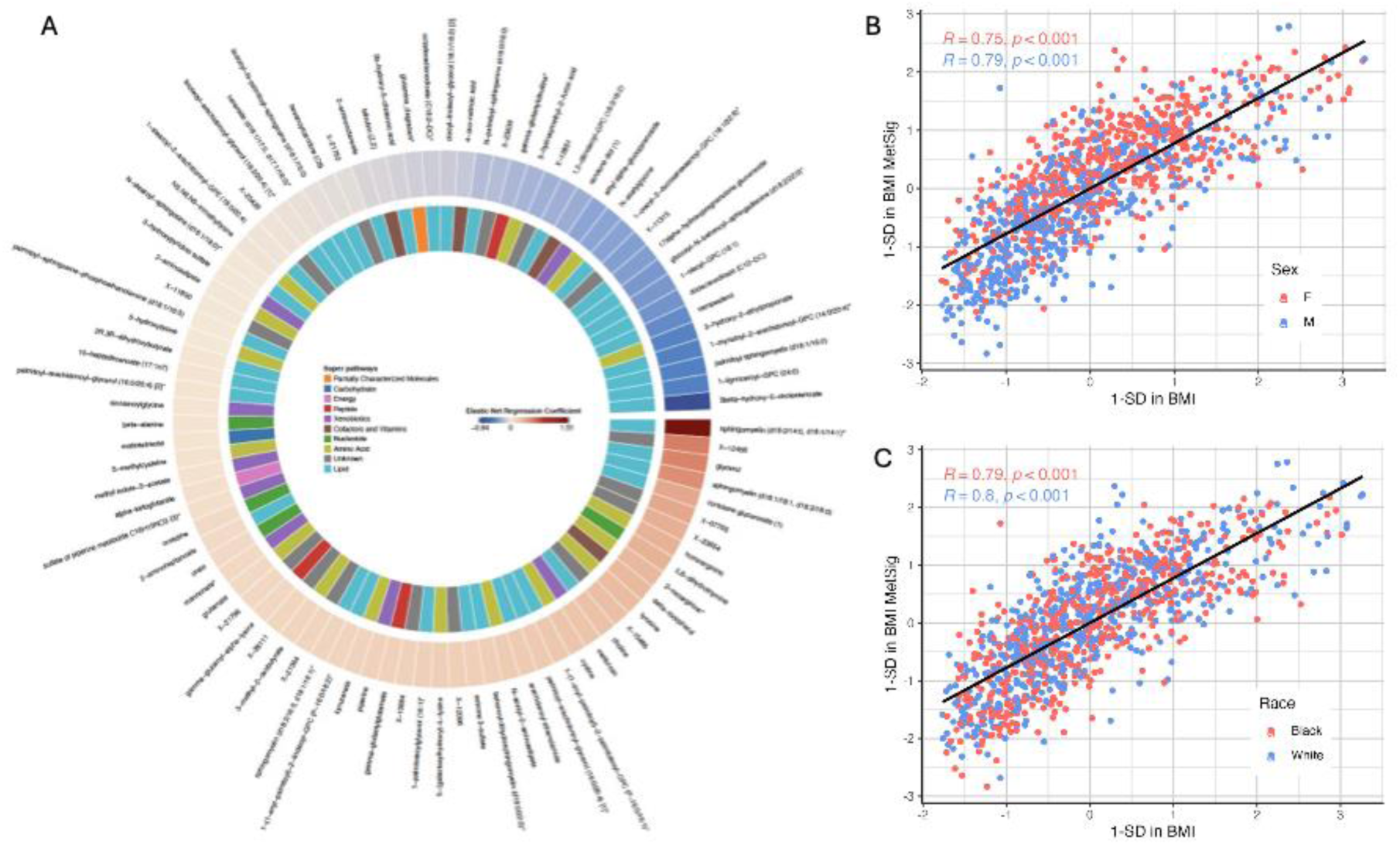
The metabolite signature of body mass index (BMI-MetSig) and its correlation with BMI by sex and race in the Southern Community Cohort Study (SCCS) **A**, The 94 metabolites selected by elastic net regression in the SCCS. **B**, Spearman correlation between metabolite signature (BMI-MetSig) and BMI in the SCCS by sex. **C**, Spearman correlation between the BMI-MetSig and BMI in the SCCS by race. Abbreviations: SD, standard deviation; F, female; M, male; p, p-value; R, Spearman correlation coefficient.

### Associations with incident CHD and CVD risk factors in the SCCS

Per 1-SD increase in BMI-MetSig was associated with a 27 mg/dL increase in triglycerides, a 6 mg/dL increase in non-HDL-C, and a 5 mg/dL decrease in HDL-C levels (all p<0.001; **Figure 2**). Each 1-SD increase in BMI-MetSig was associated with a near 50% elevated odds of incident CHD (odds ratio [OR] = 1.48, 95% confidence intervals [CI]: 1.28-1.71; **Figure 3**) after adjustment for sociodemographic and lifestyle factors. BMI-MetSig was significantly associated with incident CHD across subgroups, including men, women, Black/African American participants, White participants, ever-smokers, never-smokers, and participants living without hypertension, diabetes, or dyslipidemia (**Figure 3**). BMI-MetSig was marginally associated with incident CHD among participants with normal weight (1.46, 95% CI: 0.99-2.18) while BMI showed a null association (0.94, 95% CI: 0.34-2.58). The associations of BMI-MetSig and BMI with incident CHD were modified by hypertension history (p=0.006 and 0.001, respectively), both were more evident among participants without hypertension (**Figure 3**).

**Fig. 2.**
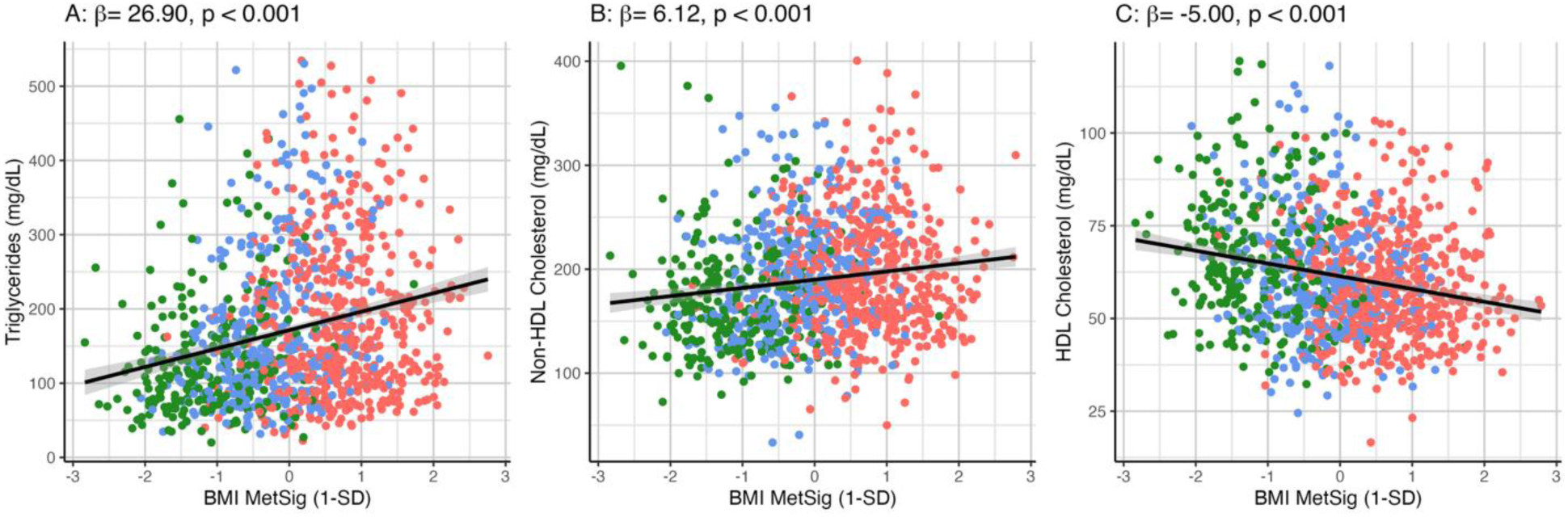
Associations of BMI-MetSig with blood lipids in the Southern Community Cohort Study (SCCS) Cross-sectional associations of per 1-SD increase in BMI-MetSig with triglycerides (**A**), non-HDL cholesterol (**B**), and HDL cholesterol (**C**) were evaluated by linear regression models, adjusting for age, sex, race, education level, annual family income, smoking status, alcohol consumption, physical activity, and diet quality. Green: normal weight; blue: overweight; red: obesity Abbreviations: BMI, body mass index; BMI-MetSig, metabolite signature of BMI; HDL, high-density lipoprotein; SD, standard deviation.

**Fig. 3.**
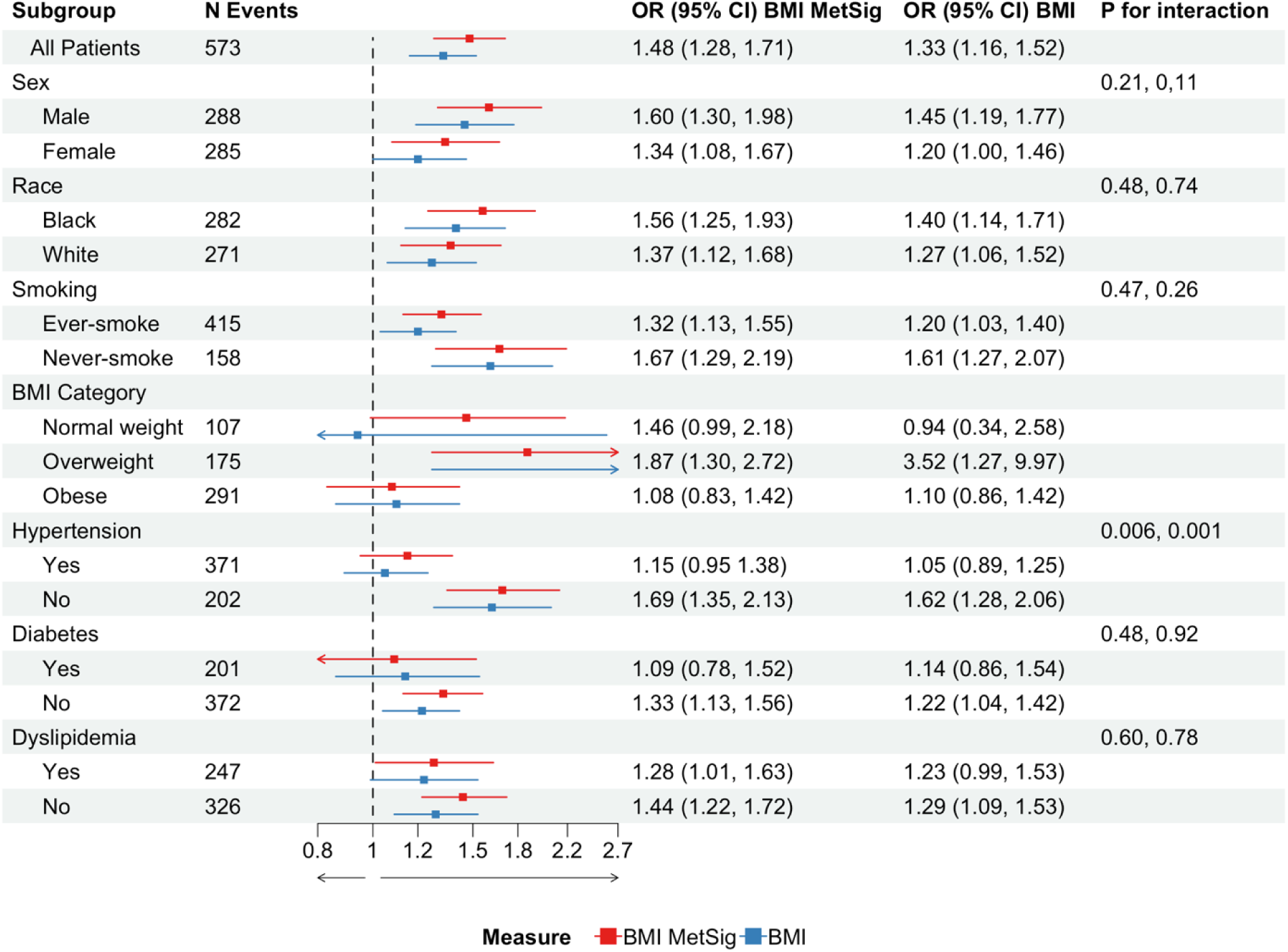
Associations of body mass index (BMI) and its metabolite signature (BMI-MetSig) with risk of incident coronary heart disease (CHD) in the Southern Community Cohort Study (SCCS)* *To acquire the odds ratio among all participants and in sex-specific and race-specific subgroups, conditional logistic regression models were used, stratified by matching pairs and adjusting for education level, annual family income, smoking status, alcohol consumption, physical activity, and diet quality. In subgroups stratified by smoking, BMI, hypertension, diabetes, and dyslipidemia, unconditional logistic regression models were used, adjusting for age at enrollment, race, sex, and covariates mentioned above. The p-values for interaction were assessed using Wald tests. Abbreviations: OR, odds ratio; 95% CI, 95% confidence interval; BMI, body mass index; BMI MetSig, metabolite signature of BMI.

Among the 94 metabolites comprising the BMI-MetSig, 33 metabolites were individually associated with incident CHD risk at FDR < 0.05, with ORs ranging from 1.16 to 1.72 for positive associations and from 0.64 to 0.86 for inverse associations (**Table S2**). Mannonate, a xenobiotic from food and plant components, had the largest adverse association (1.72 [1.51, 1.97] per 1-SD increase), followed by an unknown metabolite, two amino acids on the pathway of lysine metabolism (5-[galactosylhydroxy]-L-lysine and 5-hydroxylysine), and several other xenobiotics and lipids. On the other hand, bilirubin (Z,Z), on the pathway of hemoglobin and porphyrin metabolism, showed the second strongest inverse association after an unknown metabolite (0.77 [0.68, 0.88]), followed by metabolites on the sub-pathways of sterol, plasmalogen, and urea cycle. On the other hand, sensitivity analysis showed that all leave-one-out BMI-MetSigs had similar correlations with BMI (R ∼ 0.79) and significant associations with incident CHD (OR ∼ 1.5, p < 0.001), suggesting the BMI-MetSig results were not driven by any single metabolite (**Table S3**).

### Associations with CVD risk factors and predicted CVD risks in the GUMMY

Of the 94 metabolites comprising BMI-MetSig, 85 were found in GUMMY and used to calculate the BMI-MetSig in GUMMY using the same weights derived from the SCCS participants. A high correlation between BMI-MetSig and BMI was also observed (R = 0.75). Greater BMI-MetSig, per 1-SD, was associated with 19 mg/dL and 10 mg/dL increases in TG and non-HDL-C and 2 mL/min/1.73 m^2^ decreases in eGFR levels (all p<0.001; **Figure 4**). BMI-MetSig was associated with elevated estimated 30-year risks of CHD (1.29% higher per 1-SD greater), ASCVD (2.27%), HF (4.35%), and total CVD (3.31%) (all p<0.001; **Figure 4**).

**Fig. 4.**
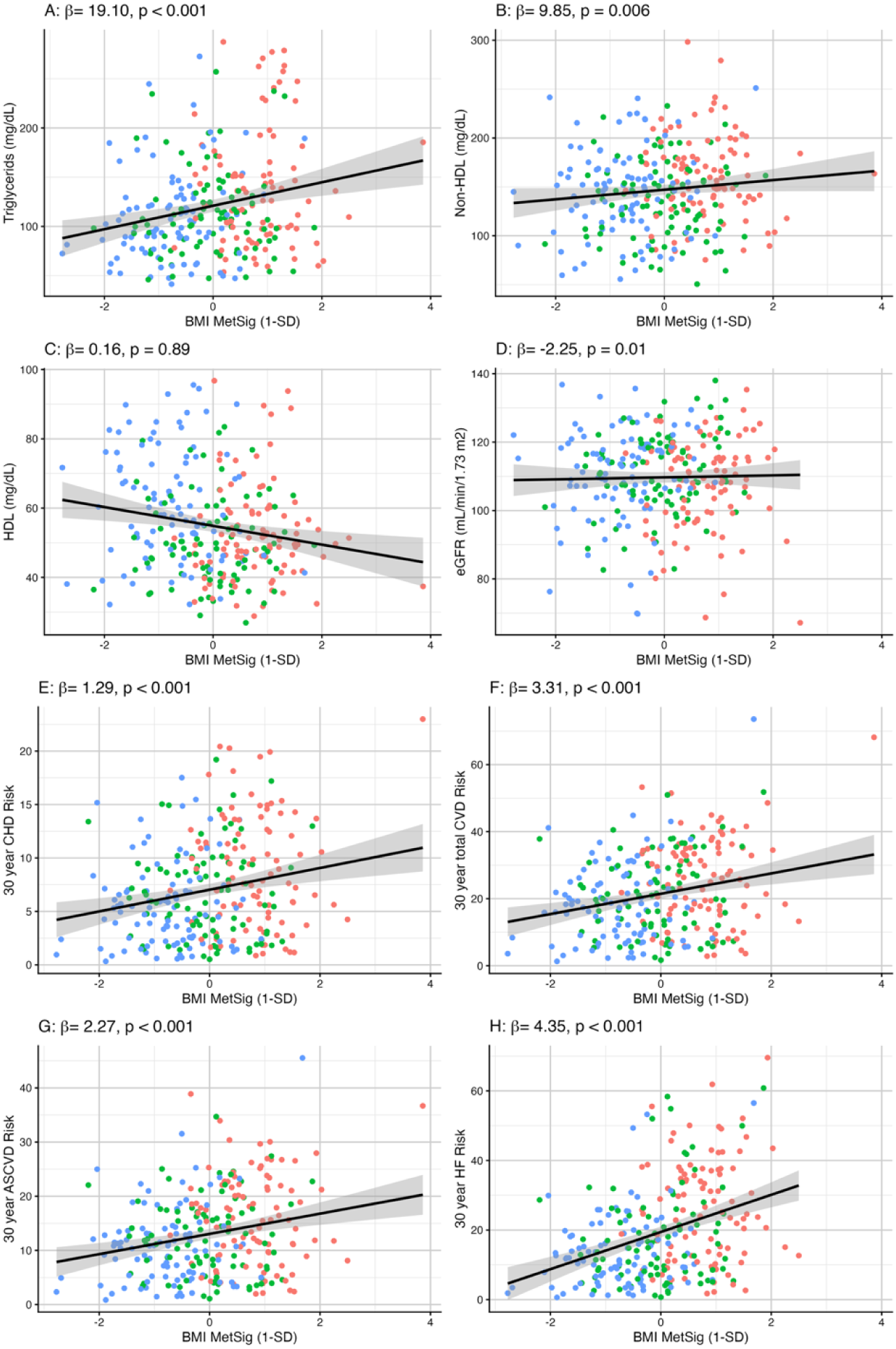
Associations of BMI-MetSig with risk factors for cardiovascular disease (CVD) and estimated 30-year CVD risks in the Gut Microbiome in Metabolic Surgery study (GUMMY)* Red: pre-metabolic surgery; green: 3 months after metabolic surgery; blue: 12 months after metabolic surgery *The associations of 1-SD increase in BMI-MetSig with risk factors and 30-year CVD risks (β regression coefficients) were evaluated by linear mixed-effects models, adjusting for age, sex, race, operation type, and histories of diabetes, hypertension, and dyslipidemia. Unadjusted simple linear regression trend lines were presented only for visualization. Abbreviations: SD, standard deviation; BMI-MetSig, the metabolite signature of body mass index

### Alternations of individual metabolites post-surgery in the GUMMY

Of the 85 metabolites comprising BMI-MetSig in the GUMMY, 17 metabolites showed significant changes within the first 3 months following MBS (**Figure 5a**), while 19 metabolites were significantly altered at 12 months post-surgery, all in the expected directions based on the BMI-MetSig weights (**Figure 5b**). The lipid choline showed significant decreases at both 3- and 12-month post-operation. At 3 months, the most significant changes involved decreases in amino acids (e.g., N-acetyl-2-aminoadipate), lipids, and xenobiotics, with bilirubin (Z,Z) and a few lipids increasing post-surgery. At 12 months, sustained decreases in amino acids (e.g.,2-oxoarginine) and xenobiotics, and increases in several lipid (e.g., lactosyl-N-palmitoyl-sphingosine) were observed. The fold changes and mixed-effects model p-values for all 85 metabolites at 3 and 12 months can be found in **Table S4**.

**Fig. 5.**
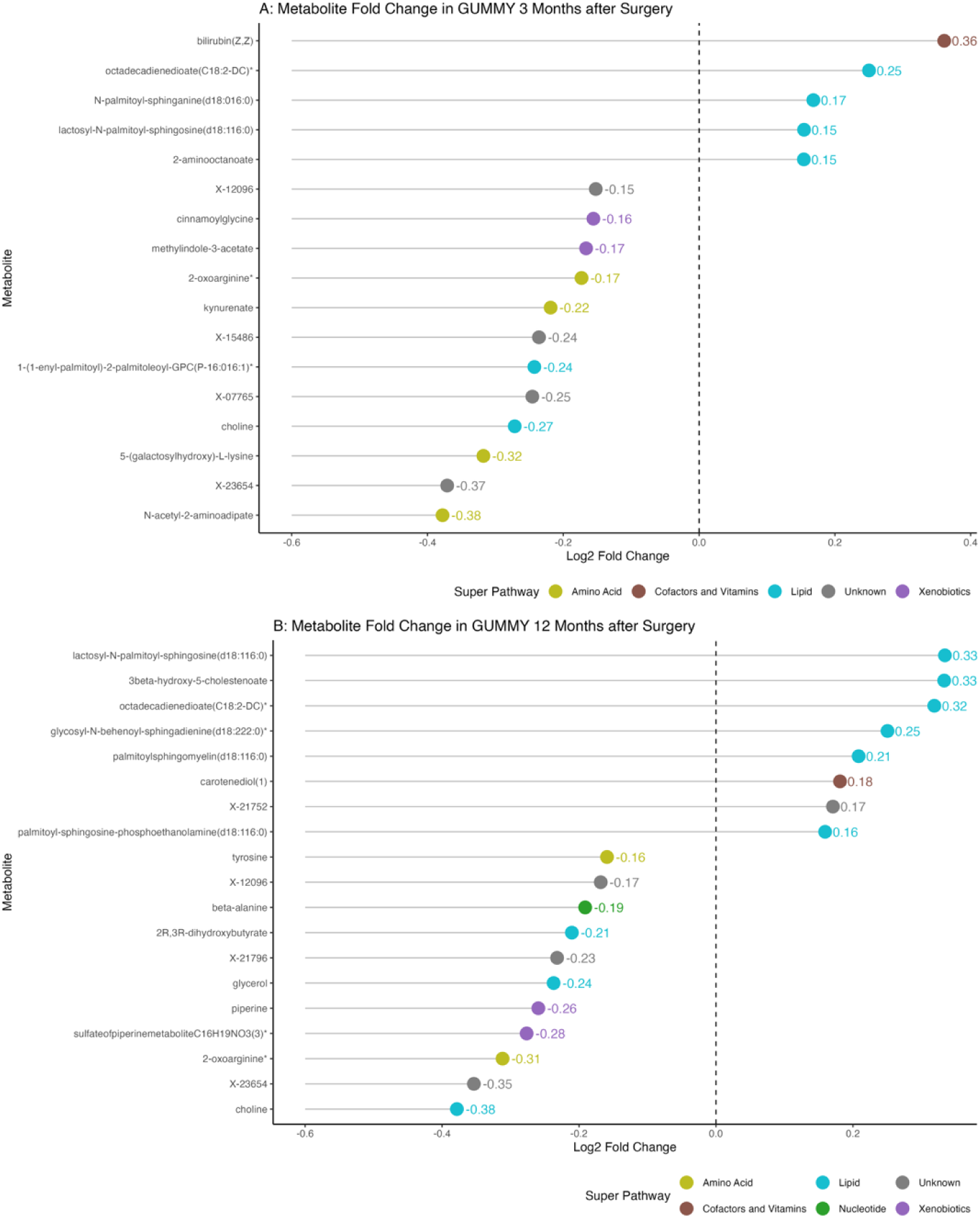
Alterations in BMI-related metabolites at 3 and 12 months after surgery in the Gut Microbiome in Metabolic Surgery study (GUMMY)* *Alterations in metabolite levels were assessed by log2 fold change and mixed effect models, adjusting for age, sex, and comorbidities. Significant changes were defined as having a false discovery rate < 0.1 and the absolute values of log2 fold change greater than 0.15.

## DISCUSSIONS

In this nested case–control study of 1,170 participants within a large cohort of Black and White Americans, we identified a BMI-MetSig consisting of 94 circulating metabolites and found it significantly associated with incident CHD, numerically stronger than BMI, particularly among individuals without obesity or previously diagnosed metabolic conditions. When applied to 95 patients undergoing MBS, the BMI-MetSig correlated strongly with BMI and predicted 30-year risks of total CVD, ASCVD, CHD, and HF. Relative concentrations of 17 (20.0%) and 19 (22.4%) metabolites in the BMI-MetSig changed significantly after surgery in the expected directions, supporting the modifiability of this signature.

### Obesity-related Metabolites

Most metabolites in the BMI-MetSig were involved in lipid or amino acid metabolism. The top-weighted metabolites were all lipids, including two sphingomyelins ([d18:2/14:0] and [d18:1/18:1]), glycerol, and cortolone glucuronide with positive weights, while 3β-hydroxy-5-cholestenoate, palmitoyl sphingomyelin (d18:1/16:0), and two glycerolphosphocholines (1-lignoceroyl-GPC[24:0] and 1-myristoyl-2-arachidonoyl-GPC [14:0/20:4]) showed negative weights. In general, sphingomyelins have been related to disorders in the circulatory system, central nervous system, endocrine system, and metabolic syndrome^25–27^. Specifically, higher levels of sphingomyelin (d18:1/18:1) in cerebrospinal fluid were associated with Alzheimer’s disease pathology^28^. Glycerol, a triglyceride backbone released from adipose tissue, has been consistently reported in BMI-related metabolic profiles^29–33^, and impaired glycerol metabolism may contribute to diabetes^34^. Cortolone glucuronide, a steroidal metabolite, has been associated with body fat distribution and metabolic syndrome^35,36^. A Mendelian randomization study has found a strong association between 1-myristoyl-2-arachidonoyl-GPC (14:0/20:4) and elevated risk of peripheral arteriosclerosis^37^. On the other hand, 3β-hydroxy-5-cholestenoate was inversely associated with BMI and mediated smoking-BMI associations in another southern US cohort^36^. 1-lignoceroyl-GPC(24:0) showed inverse associations with BMI in a Middle Eastern population^38^. Among these BMI-related lipids, glycerol was significantly associated with elevated CHD risk, while 3β-hydroxy-5-cholestenoate and 1-lignoceroyl-GPC showed significant inverse associations with incident CHD.

Obesity has been linked to several metabolic pathways, including elevated levels of branched-chain and aromatic amino acids (BCAAs and AroAAs), such as valine, leucine, isoleucine, phenylalanine, tyrosine, tryptophan, and methionine^39–41^. Consistent with prior evidence, our BMI-MetSig included metabolites from these pathways—3-methyl-2-oxobutyrate (leucine, isoleucine, valine metabolism), kynurenate (tryptophan metabolism), cystine (methionine, cysteine, SAM, and taurine metabolism), and tyrosine (tyrosine metabolism)—all previously reported to be positively associated with BMI across European, North American, and Asian populations^16,29–31,42,43^.

### BMI-MetSig and CHD Risk

Beyond their links to obesity, several metabolites within the BCAA and AroAAs pathways have also been implicated in CHD^7,44,45^. In the SCCS, we observed similar patterns—3-hydroxy-2-ethylpropionate and cystine were both associated with higher risk of incident CHD. Additionally, two metabolites involved in lysine metabolism, 5-hydroxylysine and 5-(galactosylhydroxy)-L-lysine, were significantly related to elevated CHD risk. These findings align with a prior study of 199 patients with acute coronary syndrome, in which 5-(galactosylhydroxy)-L-lysine was not only positively correlated with BMI but also elevated in patients with severe narrowing or blockage of major coronary arteries. We also identified bilirubin (Z,Z), a metabolite in the cofactors and vitamins pathway, as having the strongest inverse association with CHD incidence. This observation is consistent with previous reports suggesting potential protective effects of bilirubin on CHD and other CVD events and mortality^46–51^.

While these metabolites may help explain the biological links between obesity and CHD, our sensitivity analyses suggest that no single metabolite alone accounts for this association. Instead, the combined metabolites comprised the BMI-MetSig provides a more comprehensive reflection of obesity-related cardiometabolic risk. Indeed, BMI-MetSig consistently demonstrated numerically stronger associations with incident CHD than BMI in total participants and across nearly all subgroups—men, women, Black and White participants, ever- and never-smokers, and those without hypertension, diabetes, or dyslipidemia. Although neither BMI nor the BMI-MetSig reached statistical significance among normal-weight individuals, each 1-SD increase in the BMI-MetSig was marginally linked to 46% higher CHD risk, whereas the corresponding OR for BMI was null. This contrast suggests that the BMI-MetSig may capture cardiometabolic risk beyond body weight. Future analyses with larger sample sizes of participants within the normal BMI range may be performed to confirm this finding.

The idea that metabolic health and BMI can be discordant is not new. In the 1980s, Ruderman et al. introduced the concept of **“**metabolically unhealthy normal weight” (MUH-NW)^52^, referring to individuals with a normal BMI (< 25 kg/m²) who nevertheless exhibit insulin resistance, hyperinsulinemia, and/or hypertriglyceridemia^53^. Subsequent prospective studies have shown that 15–20% of new diabetes cases occur among people in the normal-weight range^54–56^. The new concept of clinical obesity per Lancet Commission’s definition^57,58^ also attempted to distinguish individuals at different stages of obesity and determine appropriate treatments by incorporating measures of body composition, organ function, and physical capacity. However, these assessments are resource-intensive and not always feasible in clinical practice^59^. In contrast, incorporating the BMI-MetSig alongside standard biomarkers may provide a simpler, more cost-effective, blood-based approach to reveal cardiometabolic risk, even among individuals with normal BMI and no history of metabolic diseases.

### Generalizability and Modifiability of the BMI-MetSig

While the SCCS findings demonstrate strong and consistent associations between the BMI-MetSig and incident CHD risk across participants with varied demographic backgrounds and metabolic health statuses, whether this signature applies to other study populations, including those with severe obesity, and whether it is modifiable through weight loss interventions were further evaluated among patients undergoing MBS. In the GUMMY, participant characteristics differed from those in the SCCS—patients had an average baseline BMI of 45 kg/m^2^ and were predominantly White, female, and insured for surgery. Despite these demographic and clinical differences and the absence of nine metabolites, the BMI-MetSig remained highly correlated with measured BMI in the GUMMY. Notably, the BMI-MetSig was also significantly associated with estimated 30-year risks of total CVD, ASCVD, CHD, and HF among patients before and after surgery, indicating its generalizability across study populations.

Moreover, the BMI-MetSig demonstrated modifiability by weight loss intervention, suggesting that it not only reflects cross-sectional cardiometabolic risk but also may be used to monitor cardiometabolic improvements following weight-loss interventions. Particularly, choline levels significantly decreased at both 3 and 12 months after surgery, consistent with previous studies reporting reductions in circulating choline levels after RYGB^60,61^. Another metabolite showing postoperative decrease is a lysine-pathway metabolite: acetyl-2-aminoadipate, whose precursor, 2-aminoadipic acid, has been identified as a marker of impaired glucose tolerance and future diabetes risk^62–65^. Its reduction may contribute to improved insulin sensitivity and glucose metabolism after MBS. Bilirubin (Z,Z), which had one of the strongest inverse associations with CHD in the SCCS, showed a significant postoperative increase. It reflects reduced oxidative stress or enhanced heme catabolism accompanying metabolic improvements^66–68^. Collectively, these findings underscore the modifiability of obesity-related metabolites following MBS.

### Strengths and limitations

This study evaluated obesity-related metabolites with incident CHD in a racially diverse low-income US population and then their changes after surgical weight loss. The nested case–control design within a well-characterized prospective cohort enhances temporal inference. The consistent associations across multiple demographic subgroups and metabolic disease statuses strengthen the internal validity and translational relevance of our findings. The development and application of the BMI-MetSig across two independent cohorts, one population-based and one clinical-based, further demonstrated the robustness, generalizability, and modifiability of this metabolite signature.

Meanwhile, we acknowledge several limitations in our current study. First, because of the observational nature, we could not infer causality of CHD. There may also be unmeasured confounding on the BMI-MetSig-CHD association^69^, for example, body composition (fat mass vs. fat-free mass) and body shape (waist circumference, waist-hip-ratio, and the body shape index)^70^. Nonetheless, the prospective study design and subgroup analyses by known confounders could help minimize bias. Mendelian randomization analyses or intervention studies could be employed for causal inference as a next step. Second, metabolites were profiled only once at each time point. Because metabolite levels can vary with circadian rhythms and acute factors such as fasting status and time since last meal, we could not account for within-person variability or standardize timing of specimen collection, which may have introduced measurement error. Nevertheless, the CHD case-control pairs were matched on fasting time. In addition, we acknowledge that our modest sample sizes in a few subgroups may have limited statistical power in these stratified analyses, resulting in a wide 95% CI. However, this study is still a unique addition to the literature as our study population comprised demographically diverse American individuals. Finally, the SCCS lacked data on biomarkers for glycemic control or kidney function, so we could not apply the PREVENT equations as we did in the GUMMY.

### Conclusions

In summary, we derived a BMI-MetSig in 1,170 low-income Black and White adults; both BMI and BMI-MetSig are associated with elevated CHD risk, with BMI-MetSig showing numerically stronger associations. In another clinical cohort of 95 patients undergoing MBS, the BMI-MetSig was also highly correlated with BMI and significantly associated with estimated 30-year CVD risks. Multiple metabolites comprising the BMI-MetSig changed significantly after surgery in the expected directions, highlighting modifiability of obesity-related metabolites. BMI-MetSig may provide a more comprehensive indicator of cardiometabolic health than BMI alone, and future experimental work (e.g., in cellular or animal models) could be applied to test its biomarker and therapeutic potential for weight loss.

## Data Availability

The data underlying this article can be obtained through the Southern Community Cohort Study and the Gut Microbiota in Metabolic Surgery study upon reasonable request and approval by the Data and Biospecimen Use Committees of those studies.

https://www.southerncommunitystudy.org

## Acknowledgments

The authors thank all SCCS and GUMMY participants, our clinical research coordinators, Britt T. Biesinger, MPH and Adhora Madhuri, MPH, and healthcare providers at the VUMC Weight Loss Center. The authors gratefully acknowledge Staci L. Sudenga, PhD, and Stephen Deppen, PhD, for critically reviewing the manuscript.

## AI Usage Statement

ChatGPT 5.0 was used to assist with grammar checking and language polishing for some sentences. Prompt fed to AI: “Please make this sentence more concise and clearer.”

## CRediT authorship contribution statement

**Zicheng Wang:** Formal analysis, visualization, methodology, conceptualization, writing-original draft. **Yulu Zheng:** Methodology, data curation, visualization, writing- review & editing. **Lei Wang:** Methodology, writing- review & editing. **Charles R. Flynn:** Resources, writing-review & editing. **Xiao-ou Shu:** Resources, methodology, writing- review & editing. **Qiuyin Cai:** Resources, writing- review & editing. **Deepak K. Gupta:** Resources, Writing- review & editing. **Loren Lipworth:** Resources, writing- review & editing. **Wei Zheng:** Resources, writing- review & editing. **Xinmeng Zhang:** Data curation, writing- review & editing. **You Chen:** Data curation, resources, writing- review & editing. **Jason M. Samuels:** Resources, methodology, writing-review & editing. **Danxia Yu:** Conceptualization, methodology, writing- original draft, reviews & editing, supervision, funding acquisition.

---

**Table S1.**
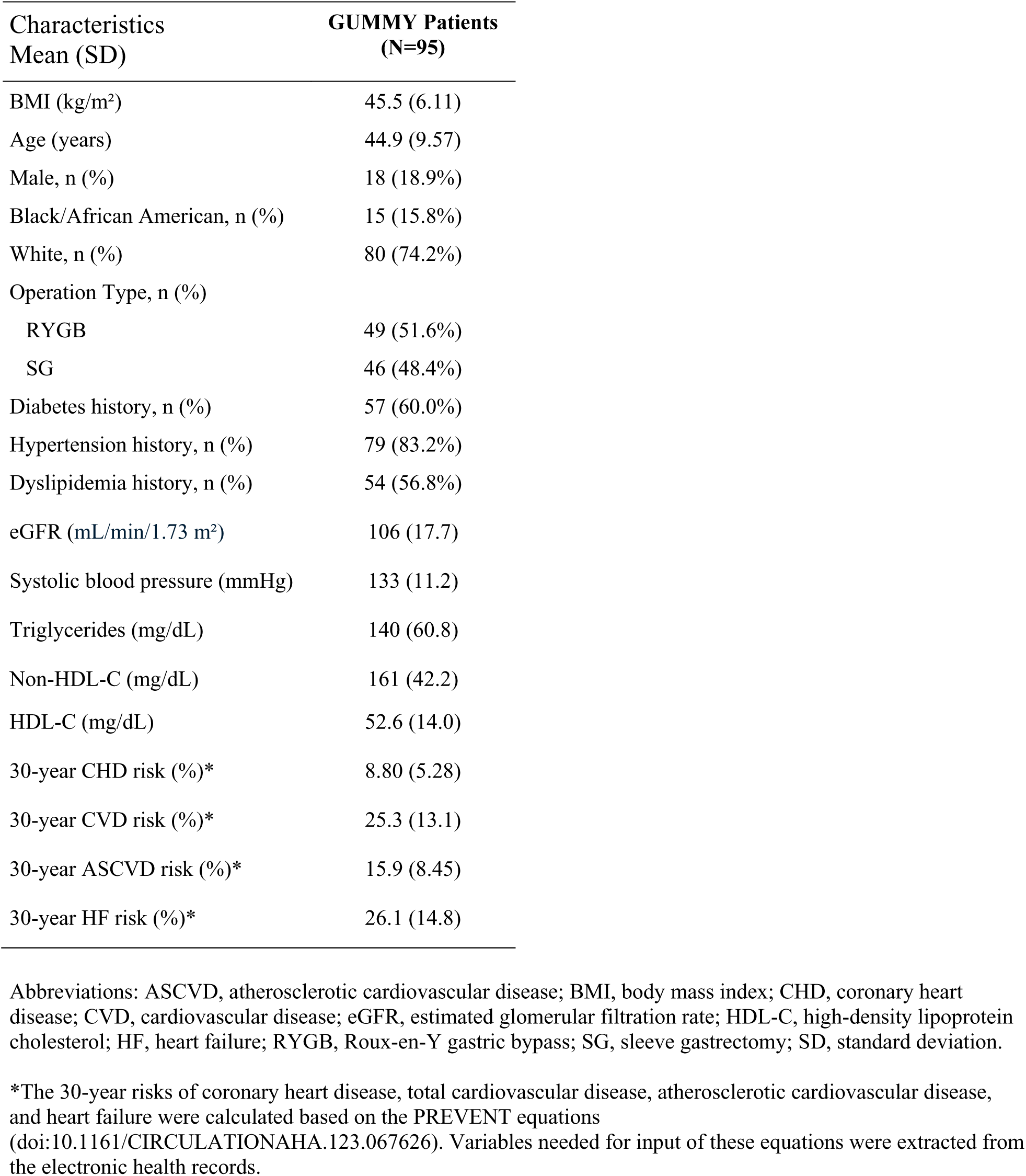
Baseline characteristics of participants in the Gut Microbiome in Metabolic Surgery study (GUMMY)

| Characteristics<br>Mean (SD) | GUMMY Patients<br>(N=95) |
| --- | --- |
| BMI (kg/m<sup>2</sup>) | 45.5 (6.11) |
| Age (years) | 44.9 (9.57) |
| Male, n (%) | 18 (18.9%) |
| Black/African American, n (%) | 15 (15.8%) |
| White, n (%) | 80 (74.2%) |
| Operation Type, n (%) |  |
| RYGB | 49 (51.6%) |
| SG | 46 (48.4%) |
| Diabetes history, n (%) | 57 (60.0%) |
| Hypertension history, n (%) | 79 (83.2%) |
| Dyslipidemia history, n (%) | 54 (56.8%) |
| eGFR (mL/min/1.73 m<sup>2</sup>) | 106 (17.7) |
| Systolic blood pressure (mmHg) | 133 (11.2) |
| Triglycerides (mg/dL) | 140 (60.8) |
| Non-HDL-C (mg/dL) | 161 (42.2) |
| HDL-C (mg/dL) | 52.6 (14.0) |
| 30-year CHD risk (%)* | 8.80 (5.28) |
| 30-year CVD risk (%)* | 25.3 (13.1) |
| 30-year ASCVD risk (%)* | 15.9 (8.45) |
| 30-year HF risk (%)* | 26.1 (14.8) |

Abbreviations: ASCVD, atherosclerotic cardiovascular disease; BMI, body mass index; CHD, coronary heart disease; CVD, cardiovascular disease; eGFR, estimated glomerular filtration rate; HDL-C, high-density lipoprotein cholesterol; HF, heart failure; RYGB, Roux-en-Y gastric bypass; SG, sleeve gastrectomy; SD, standard deviation.
\*The 30-year risks of coronary heart disease, total cardiovascular disease, atherosclerotic cardiovascular disease, and heart failure were calculated based on the PREVENT equations (doi:10.1161/CIRCULATIONAHA.123.067626). Variables needed for input of these equations were extracted from the electronic health records.

**Figure S1.**
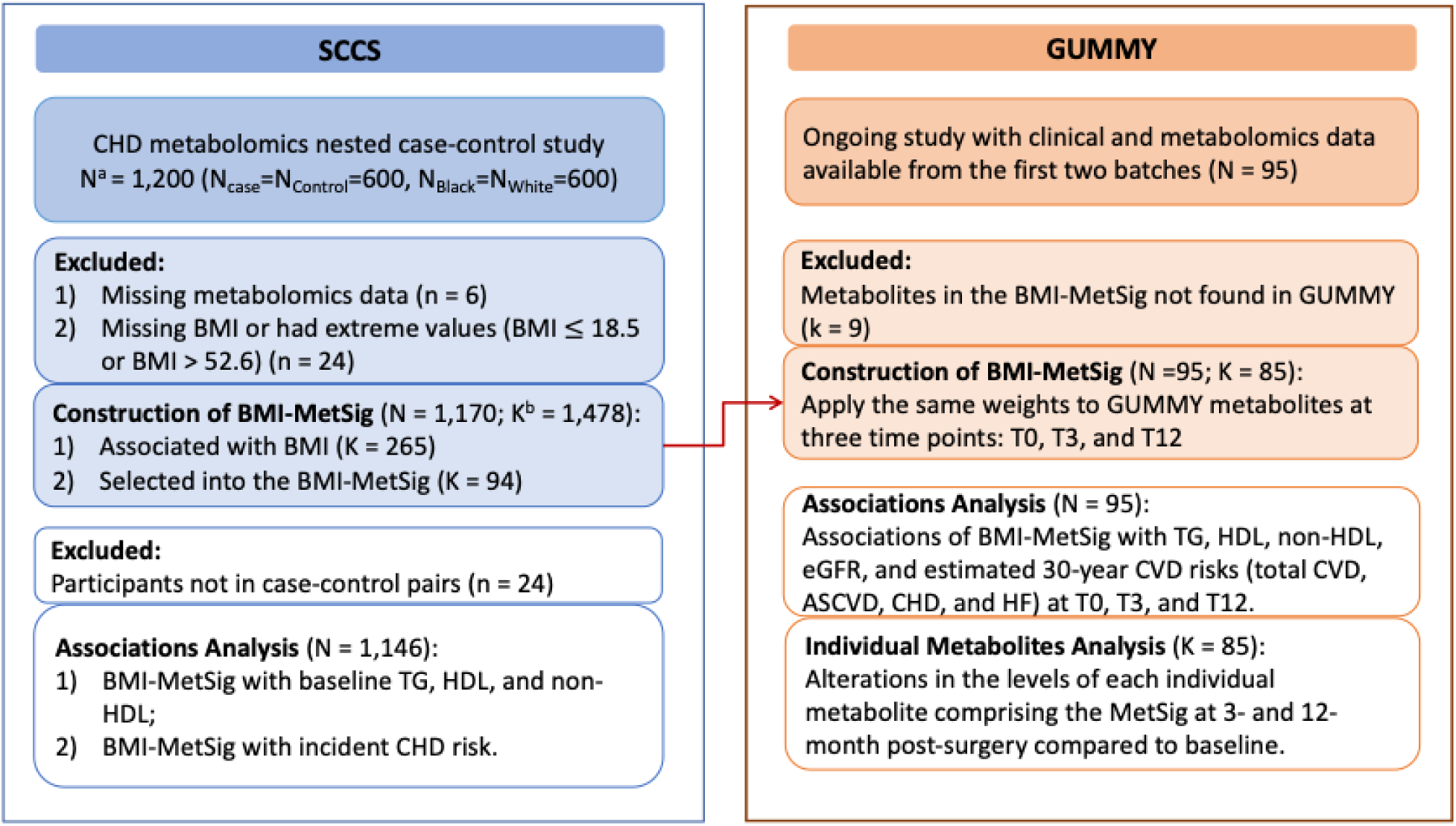
Graphical diagram of study design. ^a^N denotes number of participants; ^b^K denotes number of metabolites. Abbreviations: ASCVD, atherosclerotic cardiovascular disease; BMI, body mass index; BMI-MetSig, metabolite signature of body mass index; CHD: coronary heart disease; CVD: cardiovascular disease; GUMMY: Gut Microbiota in Metabolic Surgery Study; HDL: high-density lipoprotein; SCCS, Southern Community Cohort Study; T0: pre-surgery; T3: 3-months after surgery; T12: 12-months after surgery; TG: triglycerides.

